# Community leaders as vaccine champions and the lessons from the COVID-19 pandemic in Papua New Guinea

**DOI:** 10.64898/2026.07.21.26358216

**Authors:** Katrina Shen, Jamee Newland, Nalisa Neuendorf, Ruthy Boli-Neo, Lisa M Vallely, Agnes Mek, Anna Maalsen, Leanne J. Robinson, William Pomat, Moses Laman, Angela Kelly-Hanku

## Abstract

The lessons learned from the COVID-19 pandemic are essential for implementing substantive changes and present a significant opportunity to strengthen the resilience of health systems on both national and global scales in responding to future public health crises. It is widely acknowledged that COVID-19 vaccination represents one of the most effective strategies to contain the pandemic. Nonetheless, vaccine hesitancy continues to evoke fear and uncertainty, particularly in contexts such as Papua New Guinea (PNG), a nation characterised by one of the lowest COVID-19 vaccination rates worldwide. Insights from previous public health interventions in PNG emphasise the critical role that community leaders play in ensuring the success of such initiatives. This publication reports on a study investigating the influence of community leaders on COVID-19 vaccine uptake in PNG.

Qualitative research was carried out across seven provinces between July and November 2021, involving interviews with 131 participants, including healthcare professionals, key informants, community leaders, and primary health service clients. There was considerable support for community-led strategies, with participants advocating for awareness programs to be delivered by community leaders. The advantages of locally driven approaches include the trust communities place in their leaders and shared lived experiences, which enable them to address their specific needs effectively while respecting and building upon local beliefs and customs. The effectiveness of place-based vaccine champions was also endorsed, recognising the importance of collaborations between health systems, governments, and community leaders to promote COVID-19 vaccination.

These findings underscore the importance of sustained support and ‘bottom-up’ strategies to empower community leaders in fostering community ownership of health promotion initiatives and building resilient health systems, not only for COVID-19 vaccine distribution but also for future health interventions.

## Introduction

Vaccines are frequently heralded as one of the “greatest achievements of public health” (1), yet this view is far from universal and does not automatically translate into widespread, sustained vaccine coverage. Vaccine acceptance, and its uptake, occurs along a continuum and is influenced by the complex interplay of social, cultural, emotional and psychological factors (2). It is also affected by histories of colonization and past experiences of vaccination, as well as religion, and as evident in recent years, social media, and misinformation (3, 4). As vaccines are introduced into different cultural contexts, vaccine hesitancy and acceptance within communities are shaped by local understandings, which in turn are shaped by social, cultural, religious and other belief systems (5), and as they do, they transform the logics of them.

Caused by severe acute respiratory syndrome coronavirus 2 (SARS-CoV-2), COVID-19 was declared an international public health emergency by the World Health Organization (WHO) in January 2020 and a pandemic in March 2020 (6). The development of COVID-19 vaccines by 2021 played a crucial role in mitigating the spread and severity of COVID-19 (7). Alongside the pandemic biopolitical troubles, ‘immunopolitics’, emerged on an unprecedented scale, highlighting the self-defeating turn that biopolitics can take (8). The introduction of the COVID-19 vaccine has been heralded as the most controversial vaccination program in recent history (9). The expedited development of COVID-19 vaccines and their rollout across the globe, often facilitated by high-income nations and development partners, intensified public concerns about their safety and efficacy.

COVID-19 vaccine mandates for travel and employment were also believed to be strongly influenced by coercion and restrictions on autonomy and freedom, thereby further heightening mistrust of governments (10). In this context, anti-vaccination campaigners were readily able to exploit deeply rooted fears surrounding adverse side effects, expedited development, and the necessity of multiple doses (11). Amidst this unprecedented global crisis, the pressing need to achieve ‘herd immunity’ relied on countries’ ability to overcome vaccine hesitancy; a complex and multifactorial phenomenon defined by WHO as the “delay in acceptance or refusal of vaccines despite availability of vaccine service” (12). Despite relatively successful vaccine rollouts in some Western Pacific countries, such as Fiji, Indonesia, and the Cook Islands (13), Papua New Guinea (PNG) recorded one of the lowest COVID-19 vaccine uptake rates globally (14).

Located in the Southwestern Pacific, PNG is a sovereign nation with a population of over 11 million people, of which 87% live in rural and remote areas (15). Provided by a mix of government and church-run services, PNG’s decentralised health system, in which routine vaccination is distributed, relies on a system that was already “systematically stretched” (16) prior to the pandemic with limited resources and insufficient funding, and ongoing workforce shortages in services.

Once polio-free for 18 years, PNG’s outbreak of vaccine-derived polio in 2018 was the result of long-term low routine vaccination coverage and a lack of supplementary vaccination activities (17). The re-emergence of polio again in 2025 speaks to ongoing challenges in immunization in PNG. The national vaccination program is undermined by underutilisation of health services, stockouts from financial and logistical issues, inadequate provision of education, and vaccine hesitancy (18, 19). While research on human papilloma virus (HPV) undertaken in PNG prior to COVID-19 reported high vaccine acceptability, in at least one rural area, vaccine hesitancy was strong for all immunizations, with clear links to the history of colonization (20). In retrospect, there were significant warning signs of what was to come with vaccine hesitancy and outright rejection of the COVID-19 vaccine. Unlike all other vaccinations, COVID-19 was the only vaccine approved in PNG for those 18 years and older. PNG, like many Pacific nations, had never introduced a vaccine for adults. Thus, the rollout was met with public scepticism compounded by distrust of government interventions linked to PNG’s colonial histories, and the process of gaining informed consent with discussing risks was also reported to generate hesitancy with some individuals questioning if signing the consent form involved release of liability (21, 22).

Drawing on the testimonies of prominent national religious and political leaders, such as Catholic Cardinal Sir John Ribat and Prime Minister James Marape, PNG’s COVID-19 vaccine campaign began in May 2021 with the ‘*Sleeves Up*’ campaign. Despite the use of national leaders in the campaigns, vaccine ambivalence and hesitancy persisted, and in some cases, even intensified. Anti-vaccination protests, threats against and attacks on healthcare workers (HCWs) involved in both COVID-19 and routine immunization services forced vaccination programs and routine services to be paused or scaled back in various parts of the country, including, but not only, in Morobe and New Ireland Provinces and parts of the Highlands (23–25).

Community discourses circulating in PNG included a belief that vaccination was unnecessary as COVID-19 was imaginary with fears of side effects such as death, infertility and changes to deoxyribonucleic acid (DNA), as well as narratives of vaccines’ having microchips to sign individuals with the ‘mark of the beast’ were a means to achieve government control (26, 27). Our research on COVID-19 has argued that vaccine hesitancy went further. COVID-19 was essentially viewed as a foreign disease, and the vaccine was either irrelevant or a threat to national sovereignty and Christian worldviews. Locally resonant beliefs and opposition to the vaccine in PNG were shaped by the convergence of global conspiracies, colonial legacies of medicine in PNG and particular Christian beliefs (28). Long before COVID-19, others have noted in PNG that scientific and globalized facts about other viruses, such as HIV, have not automatically displaced local logics around illness causality and healing practices (29–32). It is therefore important not to simply dismiss these beliefs as illogical, rather, the local contexts into which biomedical interventions are introduced must be understood. It is unsurprising, then, that such discourses emerged in PNG, significantly affecting the vaccination uptake. Reliant on donations of vaccines through COVAX, Australia’s ‘donation’ of AstraZeneca to PNG, when there were growing concerns about its safety in Australia and elsewhere, further fuelled resistance and ongoing conversations about safety concerns and potential adverse effects within communities and on social media (22, 33). As a consequence, by the end of March 2023, only 4.2% of PNG’s population had received at least one COVID-19 vaccine dose, and the majority of these were HCWs and other essential workers on the frontline (34), and despite numerous campaigns, COVID-19 vaccine confidence hasn’t built over time, but rather the sentiment has stagnated (18, 35–38).

In the latter half of those early pandemic years, a growing realisation emerged that national campaigns were ineffective, and local solutions were needed. Moreover, there was an acknowledgment that coordination with local leadership had been missing. As a consequence, partnerships between non-government organisations and Provincial Health Authorities (PHAs) were formed to train and support local community leaders to become advocates for the COVID-19 vaccine in their respective communities (39). This was reported to be more successful, with community leaders describing being empowered to be vaccinated against COVID-19 and vaccine awareness in communities strengthened when delivered by trusted and influential community leaders (40). Other countries also engaged local leaders in COVID-19 vaccination efforts, as they had in other immunization programs in the years preceding the pandemic.

For more than a decade (see for example, 41) there has an important and growing body of research from low- and middle-income countries as well as among culturally, linguistically and other marginalised populations in high income settings that explores the role of and effectiveness of community leaders, students, parents and others as vaccine champions (“VaxChamps”) (as well as their training) to improve vaccination coverage. This has included work that relates to routine immunizations (42–44) and, in more recent years, HPV (45–47) and COVID-19 (43, 48–51). These studies all speak to the various types of community members who can be and are being vaccine champions, including students, parents, religious leaders, and cultural leaders, for example, often with very different roles. Across these different contexts, it is clear that often vaccine champions meet local needs others do not, for example, address misinformation (48, 51), create faith-based messaging (48, 51), build trust between authorities and their communities (42, 46, 50, 51) and empower their communities to make informed decisions about vaccination (43, 49, 50).

In this paper, we appraise the role and influence of community leaders on COVID-19 vaccine acceptability and uptake in PNG. Data is drawn from a multi-site mixed-method study exploring the impact of COVID-19 on primary health services and public health infectious disease programs in PNG (28, 52–54). At the time the study was commissioned, there was no COVID-19 vaccine, and therefore not designed to study local leaders as vaccine champions. However, local leaders are *always* central to social life in PNG and as such, by the time data collection started, COVID-19 vaccination was available, and these issues emerged in the interviews. By understanding the role of local leaders as vaccine champions in this setting, we underscore the critical role they play, and can play, in future strategies for health system resilience for pandemic preparedness and routine immunization.

### Methodology

#### Participants, study sites and data collection

The study was conducted in eight research sites across seven provinces of PNG: Port Moresby (NCD), Goroka (Eastern Highlands Province), Daru and Kiunga (Western Province), Lae (Morobe Province), Kokopo (East New Britain Province), Madang (Madang Province), and Mount Hagen (Western Highlands Province). These provinces were selected because they were deemed high-risk for COVID-19, and specific sites were selected in consultation with key collaborators with whom the research team had relationships, to ensure HIV, tuberculosis, malaria programs, and maternal and child services were represented in the study design.

Participants were recruited via snowball sampling (55). Healthcare workers (HCWs) and key informants helped facilitate contact between potential study participants and the research team. Fieldwork was conducted during PNG’s third and largest wave of COVID-19, and during the nationwide vaccine rollout from July 2021 to November 2021, by nine trained and experienced social researchers from the PNG Institute of Medical Research (PNGIMR). All team members involved in data collection were highly experienced, and trained Papua New Guinean researchers with extensive prior experience in health system research and the use of qualitative data collection tools.

Semi-structured interviews were conducted in English or Tok Pisin, following informed consent, with 131 participants from five cohorts: community leaders, key informants working within health departments, HCWs, and clients who were engaged or disengaged from primary health services (Table 1). Interviews explored participants’ knowledge and experiences of COVID-19’s impact on health systems, particularly public and primary health services. Interview questions were guided by six building blocks— health service delivery, healthcare workforces, essential medicines and medical products, health information systems, finance, and leadership and government from the WHO Building Blocks conceptual framework, which was expanded to include a seventh block of ‘people and community’ in the primary study (52, 56, 57). This places people and community at the centre of health system strengthening and overcomes the limitations of the original WHO framework by delving into not only the factors affecting the development of health interventions, but also the factors defining how such interventions are received by communities (57, 58). Written informed consent was obtained from all participants prior to participation.

**Table 1.**
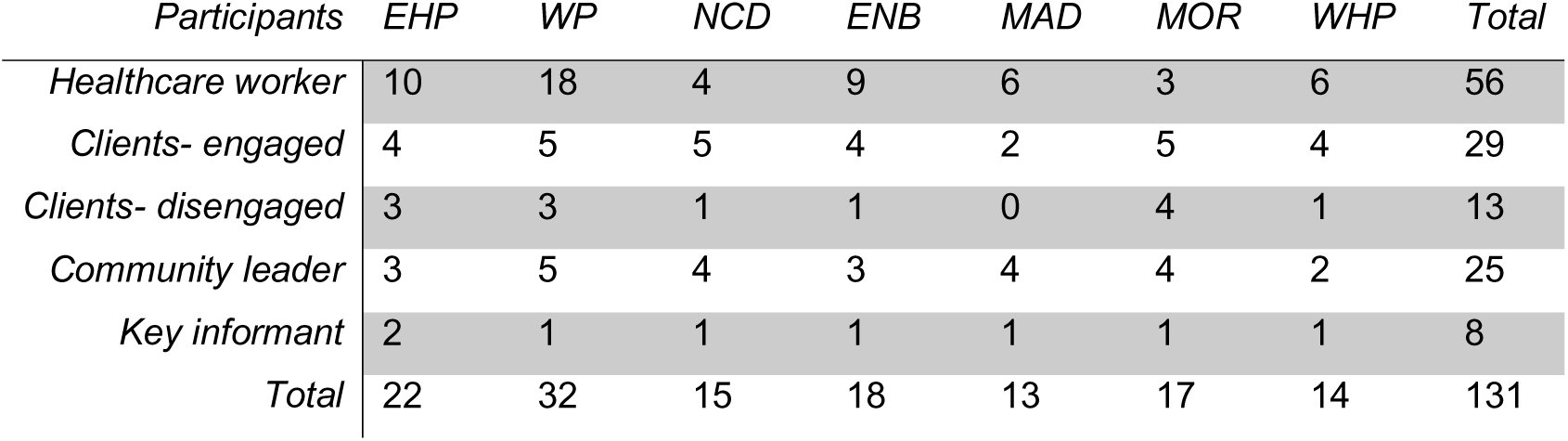
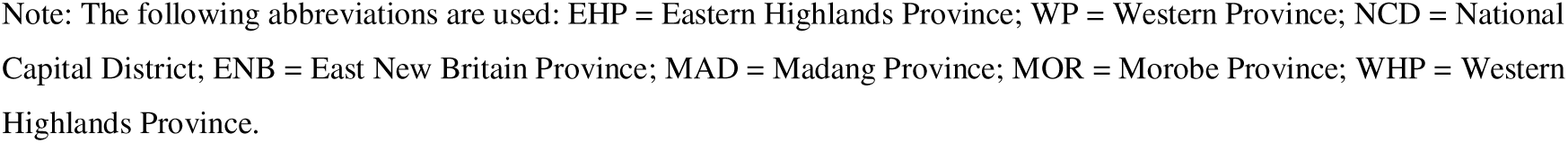
Participant numbers by province and cohort.

| *Participants* | *EHP* | *WP* | *NCD* | *ENB* | *MAD* | *MOR* | *WHP* | *Total* |
| --- | --- | --- | --- | --- | --- | --- | --- | --- |
| *Healthcare worker* | 10 | 18 | 4 | 9 | 6 | 3 | 6 | 56 |
| *Clients- engaged* | 4 | 5 | 5 | 4 | 2 | 5 | 4 | 29 |
| *Clients- disengaged* | 3 | 3 | 1 | 1 | 0 | 4 | 1 | 13 |
| *Community leader* | 3 | 5 | 4 | 3 | 4 | 4 | 2 | 25 |
| *Key informant* | 2 | 1 | 1 | 1 | 1 | 1 | 1 | 8 |
| *Total* | 22 | 32 | 15 | 18 | 13 | 17 | 14 | 131 |

Note: The following abbreviations are used: EHP = Eastern Highlands Province; WP = Western Province; NCD = National Capital District; ENB = East New Britain Province; MAD = Madang Province; MOR = Morobe Province; WHP = Western Highlands Province.

#### Data analysis and management

All interviews were transcribed verbatim and, where necessary, translated from Tok Pisin to English by trained social researchers from PNGIMR. All data has been de-identified, and pseudonyms have been assigned to maintain participants’ privacy and anonymity. The qualitative data management software NVivo version 12 (QSR International Pty Ltd) was used to store, manage, and code interview data. Coding and analysis by the lead author used deductive and inductive thematic analysis (59). Code books were produced for each key domain: 1) knowledge, attitudes, and practices of community leaders, and 2) leader positionality, which included subdomains of community trust in leaders, local understanding of community leaders, and identifying vaccine champions. The interpretation was cross-checked with the research team and co-authors, and the themes constructed by the authors underwent several rounds of revision using reflexive analysis.

#### Ethical consideration and informed consent

All participants provided informed consent in either English or Tok Pisin. Written consent was obtained from all literate participants, while witnesses signed on behalf of those who were unable to read or write. Participants also had the option to give verbal consent if they preferred. Health service clients received light refreshments. To ensure confidentiality, all study interview data and photographs with people’s faces have been anonymized. Pseudonyms were assigned to protect participants’ privacy and maintain their anonymity.

Ethics approval for this project was obtained from the PNG Institute of Medical Research Institutional Review Board (IRB #2015), the PNG Medical Research Advisory Committee (MRAC 20.35) and UNSW Human Research Ethics Committee (HC210172) and undertaken in accordance with the Australian National Health and Medical Research Council. The research team received authorisation from the seven governing PHAs to conduct the research prior to data collection.

## Results

Participants identified local ward councillors, religious leaders, elders, and business leaders as community representatives, along with others like treatment supporters and advocates for people living with disability (PLWD) who have rapport with communities. These leaders are influential in shaping community attitudes and decisions on COVID-19 vaccination. Findings highlight two themes: leaders are well-suited to oversee vaccine rollouts and investing in partnerships with them can boost vaccine acceptability and uptake.

### Community leaders are best placed to take ownership of COVID-19 vaccine rollouts

Community attitudes towards COVID-19 vaccination varied, ranging from acceptance to fear and outright opposition. Rather than relying solely on biomedical information, participants across all population cohorts and sites conveyed that vaccine uptake also depended on other social and relational factors. People’s relationships with those sharing information were viewed as critical.

#### One of their own (relationships between communities and their leaders)

Community leaders are trusted figures within local communities, sharing long-standing relationships that could be leveraged to increase COVID-19 vaccine confidence. Beetrus, a healthcare worker from the Eastern Highlands Province, said, “People in the village listen to them … they will not listen to us [healthcare workers] … Their own people, they have respect for them, and they can listen to them, so that’s the best approach.” This place-based trust depends on shared experiences, which are essential for building rapport in rural areas, as Bomali, a female community leader from the same province, stressed the importance of awareness campaigns involving local leaders, not just those who emerged from communities, but *who live in the community*, saying – “For you to reach out to their [rural settings], you need to stand with the leader who is there… they are the ones that are going to lead you back to the people… [otherwise] they will not listen to what he will say because he’s not there.”

#### Unique perspectives, understandings, and local knowledge of community leaders

Participants agreed that community leaders are best suited to lead COVID-19 vaccination campaigns, using their local knowledge to boost awareness efforts. A key issue was campaign language, which can either aid or hinder understanding. Priscilla, a church leader, suggested using visual aids instead of written materials due to low literacy rates, saying, “this needs to be thought out, planned very carefully… we are dealing with everyone… I may understand it, but down the road, where the level of education is [low, they may not understand].” In PNG’s diverse language landscape, community leaders stressed sharing messages in tok ples (local languages) rather than only Tok Pisin and English. They explained this helps people understand COVID-19 vaccination information and make informed choices. In Western Province, where research was conducted in Daru and Kiunga, vaccination campaigns used Kiwai, one of the local dialects. Local leaders fluent in their language helped improve vaccination rates. Fred, a security guard, said, “People started coming one by one,” and added that community leaders translated information into Kiwai, encouraging more to get vaccinated.

Participants also unanimously stressed the need to engage dedicated local leaders trusted by their communities to start conversations about COVID-19 vaccination and co-create material in ways which address community-specific concerns and needs. These leaders need to be more than simple messengers of biomedical information. They must be trained and skilled in interpreting such information and presenting it in ways that make sense locally, creating a synergy between sometimes competing models of understanding health and illness (31).

In a predominantly Christian country, participants spoke of how church leaders and religious institutions wield powerful influence over communities’ understanding and opinions on health matters, particularly vaccination uptake. Speaking to the need to synergise models of health beliefs, Anne, a female leader, described that the ability of religious leaders to tailor vaccine messaging around theological points could encourage Christian populations to receive COVID-19 vaccination. She shared, “It’s better for a religious man or a religious woman to travel with you people for this [vaccine awareness] …he/she can quote Bible verse…we are the image of God, and God loves us, so we should live.”

Similarly, for certain population groups who face structural, social, and economic barriers to healthcare, leaders were also reported to be significantly influential in disseminating information and shaping vaccine decision-making. Alex, a male community leader representing people living with disability, exposed how mainstream COVID-19 vaccination campaigns did not effectively reach his community, as it was ‘second-hand information.’ Instead, he emphasised the need for a door-to-door approach for information to reach people living with disability in their community settings.

### Investing in partnership with community leaders

To facilitate the integration of various models of understanding health and illness and to translate biomedical information into locally relevant ways that address community-specific needs, investing in grassroots leadership through vaccine champions is crucial. The official information produced and disseminated by the national government, often based on global campaigns from development partners including WHO, was frequently perceived as ineffective at the local level. In this context, and amid growing resistance to COVID-19 vaccination driven by the convergence of global conspiracies, cultural beliefs, apocalyptic Biblical prophecies, and the colonial legacies of medicine in PNG, the circulating information failed to address these concerns.

#### Empowering community leaders as vaccine champions

Amidst the threat of the COVID-19 pandemic, many community leaders expressed a determined interest in becoming vaccine champions for their diverse communities. They spoke of their desperate desire to improve the country’s vaccine rollout and suggested inviting local community leaders with established trust and rapport to receive the COVID-19 vaccination and advocate for wider COVID-19 vaccination uptake. This was observed to have early success in Western Province, with Peter, a key informant, noting, “If we can get COVID-19 champions, a pastor to be a champion in the community, I’m sure the flock would follow … When the leader got the vaccine, everybody in the village was standing in line [to receive the vaccine”.

The success of training and recruiting vaccine champions went beyond Western Province and the Church. Community leaders reported that receiving the first dose and not being marked with the beast or experiencing other adverse events, eased their doubts and increased their confidence to promote vaccination and counter misinformation circulating via social media. Mutual trust, collective responsibility, and reciprocity are core to PNG culture, with initiatives like bilums fostering empowerment and a sense of belonging among women. Bomali, a business leader from Eastern Highlands, encouraged her community of bilum weavers to vaccinate after becoming a vaccine champion. She told them, “there is so [much] garbage on social media… and it makes everybody very afraid…I got vaccinated and nothing happened, you need to take this. When I influence them, they will follow… I had to keep my weavers safe because I needed them, they need me, and I need them.”

Other community leaders had not yet been vaccinated against COVID-19 but expressed a desire for accurate, reliable information to help them and their communities make informed decisions. Darnel, a local ward councillor from the Western Highlands Province, also spoke of how information was not filtering down, explaining “The [vaccine] message is not conveyed properly from the Parliament, so when it reaches our level, it confuses us … We have to be engaged [in] awareness and then report back to them, then from there you will see people looking after themselves. Anyway, there is no such thing”. Similarly, Kia, a male peer educator involved in HIV and TB services in Morobe Province, emphasised the need for training to empower community leaders as trusted COVID-19 vaccination interpreters. This is crucial for key populations like female sex workers and transgender individuals, who face higher HIV risks and socioeconomic barriers. Kia stated, “Teach us the basics of COVID so we can provide accurate information in our communities." Others discussed navigating COVID-19 vaccination conversations and addressing concerns related to HIV, TB, disability, religious beliefs, and family traditions.

#### The limits of community leaders as vaccine champions

Some community leaders endorsed the scientific benefits of vaccination, but most expressed doubts, influenced by beliefs in traditional remedies, fears of microchips, and apocalyptic ideas, which sometimes hinder vaccine acceptance. Kerapeng noted that community leaders like pastors can both promote and obstruct vaccination efforts, with misinformation linking the vaccine to the mark of the devil - ’666’ - undermining outreach. Jerry, a District Health Board Chairman from a rural area in East New Britain Province, spoke about the belief in “PNG natural immunity,” which combines faith in God’s natural protection with the idea that the virus originated outside PNG. Along with reports of side effects, people, including himself, choose to rely on traditional disease prevention methods intertwined with local Christian beliefs because “God created us with natural immunity”.

Many community leaders in this study felt uncertain about the vaccine, which led most to remain neutral, neither supporting nor opposing vaccination. The lack of reliable, accessible information about COVID-19 vaccination hindered their ability to decide, limiting their capacity to become vaccine advocates. Additionally, they felt frustrated by their neutrality, as they believed they could not effectively fulfil their leadership roles. Colin, a ward councillor, expressed his frustration and concern about his ability to gather and share such information - “They [governments] just withheld everything to themselves, and they wanted to do it [awareness] themselves… Not that [giving information about vaccine to community] because what if I give them false information?”

Illustrative of a minority perspective, a few community leaders in this study spoke of actively advocating against COVID-19 vaccination. For example, Vince, a peer educator for people living with TB from the National Capital District, was concerned about the deprivation of autonomy because of COVID-19 vaccination mandates for employment and air travel and described mobilising his community to stand against the vaccine rollout, viewing it as coercive. He emphasised, “They are not animals [that] you take, chain, and start injecting… We’ll gather our people together… It’s going to be easy, and we will definitely oppose it.”

#### The need for a unified approach in collaboration

Reflecting on the precarious balance between desire for protection against COVID-19 infection and fear of the vaccine developed to protect against it, participants noted the limited success of PNG’s vaccine campaign. Critically, they acknowledged that the influence of vaccine champions will remain limited if fear and uncertainty continue to dominate and remain unaddressed and conflicting discourses amongst leaders remain unabated. Philip, a treatment supporter for people living with TB, stressed the need for consistent messaging about COVID-19 vaccination, ensuring that all leaders are adept at explaining COVID-19 vaccine efficacy and safety profiles in locally relevant, relatable ways for non-experts. For Philip, who was yet to be vaccinated, encouragement from vaccine champions was not and will never be enough because “We have to stand on one side, on our side of the story, we have to be confident in what we are talking about … We are confusing everybody”.

Building on insights from participants like Philip about the need for consistent messaging and skilled vaccine champions, others emphasised a unified approach focused on trust-building. Rashida, a nurse from Western Highlands, called for community-led efforts that prioritise listening, relationship-building, and addressing questions through conversations supported by skilled vaccine champions who can translate scientific facts into relatable language. She explained, “People are in fear. But if only we could gather all those people, tribe by tribe, get their views, listen to them, stay with them, and answer questions in a simple way … They could spend time sitting and answering the questions, which would clear their minds.” Morris, a nurse from Morobe Province, added, “We also brought community leaders to be with their people, talked, answered questions, and provided feedback… Helping break down barriers.” One participant praised ongoing partnerships between health coordinators and community leaders to address the needs of people with disabilities, noting these conversations help reduce fears. However, vaccine distribution logistics limited uptake, as Alex, a community leader from Morobe, said, “There was a lot of fear, so she came, and we held a session.” In this case, the vaccine had not yet been distributed.

## Discussion

Community leaders in PNG have long been recognised as important and influential stakeholders, including in health programs (60, 61) and the lessons learnt from the COVID-19 pandemic reinforce the need for a bottom-up, coordinated multi-sectoral approach to improve health system preparedness for future health crises. In our study, two key themes emerged. The first examines the capacity, role, and reach of community leaders, and the second considers the benefits of investing in partnerships with them to build a resilient health system.

Communities’ relationships with their leaders are founded on a shared understanding of social, cultural, and religious life, and, through this, trust. Community leaders are uniquely positioned to influence how health programs are implemented and ultimately accepted, including vaccination. This was not utilised in PNG’s early response to COVID-19. The delivery of COVID-19 vaccination information by trusted community leaders is reinforced by other community health initiatives in PNG, including responses to cholera outbreaks and improvements in access to maternal and child health services (62, 63). While trust and shared local understanding for participants in this study were constructed as two separate domains, in reality, they intersect, underpinning community leaders’ relationships with their communities; one cannot exist without the other. Community leaders spoke of the central trust they have in their respective communities, developed through established and maintained long-standing rapport, familiarity, and shared lived experiences; this places them in an important position to localise global and national health programs in situ.

Participants reported that health information about COVID-19 vaccination should be delivered by community leaders who reflect and represent not only cultural groups but also other specific populations, such as people with disabilities and other key population groups. Due to shared experiences, community leaders are best placed to recognise and understand the various barriers to COVID-19 vaccine uptake, and to assist in the co-design of local solutions unique to their respective communities. As part of this approach, participants spoke of the need to move beyond Tok Pisin and English, and translate messaging into local languages, a finding consistent with other linguistically diverse settings (64, 65). Community leaders also advocated for information to be made more accessible to communities using lay language and visual messaging, especially important in a country with low levels of literacy (66, 67). Social media was commonly reported as a source of information, and many participants, both individuals and community leaders, were hesitant to get vaccinated after hearing narratives on Facebook and Whatsapp, amidst conflicting information posted by government officials and contexts where social media fact-checking is infrequent (18, 21, 22). Religious leaders also suggested that messaging would be better received by Christian’s if theological arguments were delivered by the clergy to encourage COVID-19 vaccine uptake. Elsewhere, the value of incorporating religious messaging into COVID-19 messaging was reported (48). Although the early ‘Sleeves Up’ campaign utilised testimonies from prominent national leaders, its success in increasing COVID-19 vaccine acceptability was limited by the minimal engagement of local community leaders who spoke to people in their communities (18, 27).

Key to community leaders’ ability to promote COVID-19 vaccination was the strong, direct relationship between their own understanding of the vaccines and what they promoted or did not. The values, beliefs and concerns expressed by community leaders suggest that their stance towards COVID-19 vaccination cannot be classified as simply vaccine acceptance or refusal but rather represent a continuum, a finding consistent with theories on vaccine hesitancy (2, 68, 69). This continuum appeared heavily influenced by the precarious balance between fear and desire for protection amidst the prominence of misinformation. While some narratives, such as those centred on fears of side effects, were echoed globally, others were specific to PNG, such as the belief of ‘local immunity’, elsewhere, and these were echoed in the discourses of both community leaders and members (2, 70, 71).

The lack of priority given to strengthening and fostering community leaders’ knowledge of COVID-19 and the biomedical approaches used to prevent infection meant that such leaders were frequently unable to address the complex interplay of fear, sociocultural beliefs, and distrust of authorities underlying vaccine hesitancy in their communities, sometimes themselves too. The introduction of new biomedical technologies such as COVID-19 vaccination requires careful consideration of local milieu, especially given the historical distrust of vaccination in PNG (2, 5, 72). Engaging with communities, meeting them were they are at, is vital to address low vaccination coverage and equipping leaders with simplified, accurate information on COVID-19 vaccination so that they are empowered, skilled and confident to guide, local narratives (73, 74). But more than this, they can resist and re-shape the harmful, counter heath narratives that arise.

Health system resilience relies on effective governance and engagement of leadership systems in communities, especially in settings limited by funding, workforce, and resource constraints such as PNG. Community leaders and HCWs advocated to strengthen partnerships between health authorities, community leaders and governments to expand the capacity, role and reach of leaders’ influence on COVID-19 vaccine acceptability. The influence of vaccine champions cannot exist in isolation if fear and uncertainty continue to dominate, especially amidst the conflicting advice of various information sources such as churches, governments, and prominent individuals. As in other settings (46, 48, 50), in this study/in other PNG research campaigns were more successful in increasing wider uptake of vaccination when co-designed, shared and endorsed by community leaders. Thus, community-oriented approaches should involve leaders from all communities and provinces in decision-making and provide them with the opportunity to voice their local knowledge and concerns to best support health systems.

When provided with training on COVID-19 vaccination, community leaders could assume the role of vaccine champions to act as local role models. After being vaccinated, they could encourage people to make an informed decision and influence wider community vaccine uptake. Such strategies were successful in Daru, Western Province, for example, which may be a result of early collaborations that trained and recruited local leaders from a range of communities, such as church leaders, people with disability advocates and TB treatment supporters (75). By harnessing the trust and local understanding of place-based vaccine champions, the Western Province rollout was better equipped to attend to local beliefs and concerns. Furthermore, the findings support implementing grassroots leadership in public health programs where peer educators, HCWs, and other community volunteers are also empowered to become vaccine champions in order to act as a catalyst for change in sustainable community ownership, a finding with locally relevant evidence in programmes to improve women’s and child health which advocates for health promotion to be led and maintained by communities (62).

Whilst the peak of the COVID-19 pandemic has passed, this study presents critical insights into future directions for Papua New Guinea’s health system and leadership in the aftermath of COVID-19 and its health, socioeconomic and political ramifications (52, 76). The compelling support for campaigns to be locally driven by community leaders speaks to the importance of engaging community leaders as part of efforts to build resilient health systems. The engagement of community leaders and consideration of local settings in PNG in future vaccine programs and emerging health crises may result in a wider and more successful outcome if grassroots leadership is implemented in targeted communities. It is also imperative to address the potential for community leaders to advocate against health interventions. Therefore, community leaders must be equipped early with information from accurate, reliable sources to counter the power of misinformation circulating among both community leaders and their communities.

The lessons learnt from PNG’s health system and leadership infrastructure’s response to COVID-19 suggest the need to involve local and traditional governance systems in future public health responses to effectively engage local communities. Local structures of village councillors and health committees have been in effect for many years, and should be further strengthened to not only lead to effective service delivery, but also build trust and long-standing partnerships to help bridge critical gaps in PNG’s health system. Community leaders and other trusted individuals can easily catalyse change, and thus should be identified early and empowered via training and revision of leadership structures, such that they are able to take on a significant role rapidly and effectively in future health interventions (52). Community leaders do not exist in isolation, and study findings support global calls for public health responses in LMICs (50, 77, 78) to be adaptable and collaborative to facilitate swifter implementation of future public health interventions and effectively respond to emerging crises.

The timing of fieldwork for the study coincided with PNG’s third and largest COVID-19 wave from July to November 2021, which impacted data collection but likely influenced community discourses about the pandemic and the biomedical approach to prevention through vaccination. Still, collected at the peak of risk offers a rare glimpse into the response to COVID-19, providing critical lessons for both pandemic preparedness and routine immunization going forward. The influence of community leaders on COVID-19 vaccination program was not a focus of the study but emerged through inductive analysis as a key issue. It was noted that the majority of participants who spoke about community leaders were community leaders themselves, HCWs or key informants, with only a few people who were clients of primary health services. PNG is a culturally diverse country, and while qualitative research does not endeavour to be representative, but rather seeks to capture local perspectives regarding the potential for leadership in COVID-19 vaccine rollouts, this study is the most comprehensive qualitative study to date, undertaken across 8 sites in 7 provinces with 131 participants. Furthermore, since COVID-19 vaccination had been available for only several months, these findings are likely to reflect community leaders’ understanding and positionality regarding COVID-19 vaccination, which in turn influences community uptake amidst the fluctuating challenges of unprecedented times. Further research is needed to explore the intricacies of the dynamics among stakeholders and communities’ responses, with more extensive data to understand the extent to which vaccine champions’ influence can translate vaccine acceptability into vaccination.

## Conclusion

These findings speak to the crucial role that community leaders who hold a trusted position in their societies can play as vaccine champions in a pandemic and in routine immunization programs. Collected at a critical time in PNG’s COVID-19 history, these findings speak to the need to invest in locally driven vaccine awareness, fostering relationships and building trust in the communicator and in what they are voicing. The means by which health information is shared, by whom, how, and where, is key to fostering this trust and building vaccination acceptability in the hope that this results in the desired behaviour change of increasing vaccination coverage. In the rush to build herd immunity during the pandemic, the process necessary to achieve it was overlooked. This study also contributes to a wider understanding of leadership in PNG, reinforcing the idea that trust is often socially situated, place-based and thus, not transferable (79–81). The lesson, moving forward, for building a resilient and healthy society is to invest in this now, both for routine immunization and to create the conditions that are favourable for responding to future pandemics when, not if, they come.

## Data Availability

The minimal data set is available on reasonable request to the Papua New Guinea Institute of Medical research via

